# Obstructive sleep apnea is associated with specific gut microbiota species and functions in the population-based Swedish CardioPulmonary bioImage Study (SCAPIS)

**DOI:** 10.1101/2022.06.10.22276241

**Authors:** Gabriel Baldanzi, Sergi Sayols-Baixeras, Jenny Theorell-Haglöw, Koen F Dekkers, Ulf Hammar, Diem Nguyen, Yi-Ting Lin, Shafqat Ahmad, Jacob Bak Holm, Henrik Bjørn Nielsen, Louise Brunkwall, Christian Benedict, Jonathan Cedernaes, Sanna Koskiniemi, Mia Phillipson, Lars Lind, Johan Sundström, Göran Bergström, Gunnar Engström, J Gustav Smith, Marju Orho-Melander, Johan Ärnlöv, Beatrice Kennedy, Eva Lindberg, Tove Fall

**Author notes:** Corresponding author: Tove Fall, EpiHubben, Uppsala University 751 85 Uppsala, Sweden. Shared senior authorship (Eva Lindberg, Tove Fall).

## Abstract

Obstructive sleep apnea (OSA) is a common sleep-related breathing disorder. In animal models, OSA has been shown to alter the gut microbiota; however, little is known about such effects in humans. Here, we used respiratory polygraphy data from 3,570 individuals aged 50–64 from the Swedish CardioPulmonary bioImage Study (SCAPIS) and deep shotgun metagenomics to identify OSA-associated gut microbiota features. We found that OSA-related hypoxia parameters were associated with 128 bacterial species, including positive associations with *Blautia obeum* and *Collinsela aerofacines*. The latter was also associated with increased systolic blood pressure. Further, the cumulative time in hypoxia was associated with nine gut microbiota metabolic pathways, including propionate production from lactate, a biomarker of hypoxia. In conclusion, in this first large-scale study on gut microbiota alterations in OSA, we found that OSA-related hypoxia is associated with specific microbiota features. Our findings can direct future research on microbiota-mediated health effects of OSA.

## Introduction

Obstructive sleep apnea (OSA) is characterized by upper airway collapse episodes during sleep resulting in complete cessation (apneas) or reduction (hypopneas) of air flow and consequent intermittent hypoxia^1^. It is estimated that OSA affects 5–36% of the adult population, depending on the country studied and the diagnostic criteria used. The increasing prevalence of OSA has been attributed to the increased longevity worldwide and the rising prevalence of obesity^2^, a well-described cause of OSA^3^. Although OSA has been prospectively associated with cardiovascular disease independent of BMI^4, 5^, the mechanisms are not yet fully elucidated^6^.

The most commonly used clinical parameter of OSA severity is the apnea-hypopnea-index (AHI), which quantifies the number of apnea and hypopnea events per hour of sleep. However, AHI does not differentiate short apnea events with mild oxygen desaturation from prolonged events with severe hypoxia^7^. To quantify the time in hypoxia during sleep, the assessment for OSA often measures the percentage of the sleep time with oxygen saturation <90% (T90). The T90 parameter is a more reliable metric of nocturnal hypoxia intensity but not always associated with AHI^8^. Lastly, the oxygen desaturation index (ODI), which quantifies the number of oxygen desaturation events per hour of sleep^9^, is considered the most suitable parameter when the sole purpose is to measure intermittent hypoxia^10^. In sum, the three parameters are complementary to each other as they capture different dimensions of OSA.

The microbiota is a complex microbial community that interacts continuously with the host^11^. Studies of animal models of OSA have found that intermittent hypoxia and intermittent airway obstruction can produce substantial changes in the gut microbiota composition^12–15^. In turn, alterations of the gut microbiota induced by OSA may partly mediate the effects of OSA on adverse health outcomes, including the development of hypertension^15–17^. Smaller studies in humans have linked OSA to the microbiota composition in the upper airways (n = 92)^18^ and in the gut (n = 113)^19^. However, these studies did not adjust for important confounders, such as diet and medications, and were limited in their taxonomic resolution of the microbiota.

To overcome the above limitations, there is a need for adequately powered investigations of the associations between OSA and the human gut microbiota, combining extensive information on confounding factors with species-level microbiota data. Here, we used a validated method suitable for population-wide screening for OSA (ApneaLink Air®, ResMed, CA, USA)^20, 21^ to investigate how key prognostic and severity features of OSA were cross-sectionally associated with the human gut microbiota, analyzed with shotgun metagenomic sequencing in up to 3,570 participants from the large population-based Swedish CardioPulmonary BioImage Study (SCAPIS). We found that all three OSA parameters were associated with lower gut microbiota richness and evenness, as measured with the Shannon index. In addition, T90 was independently associated with 59 specific bacterial species and ODI was independently associated with 97 species. We further found that the T90-associated microbiota were enriched for nine microbial metabolic pathways. One of the T90-associated species, *Collinsella aerofaciens*, was also associated with systolic blood pressure independent of BMI, the OSA parameters, and related risk factors. These findings may guide future research on the role of the gut microbiota as a potential mediator of OSA-associated morbidities.

## Results

### Descriptive statistics

After excluding the participants who reported use of continuous positive air pressure (CPAP) as treatment for OSA (n = 59), the study sample consisted of 3,175 (54% female) SCAPIS participants with valid AHI data (Fig. S1) and 3,570 (52% female) with valid T90 and ODI data. The mean age was 57.7 years. The Spearman’s correlation coefficient between the OSA parameters was 0.56 for AHI and T90; 0.92 for AHI and ODI; and 0.63 for T90 and ODI. Population characteristics are described in Table 1, as well as in Tables S1 and S2.

**Table 1.**
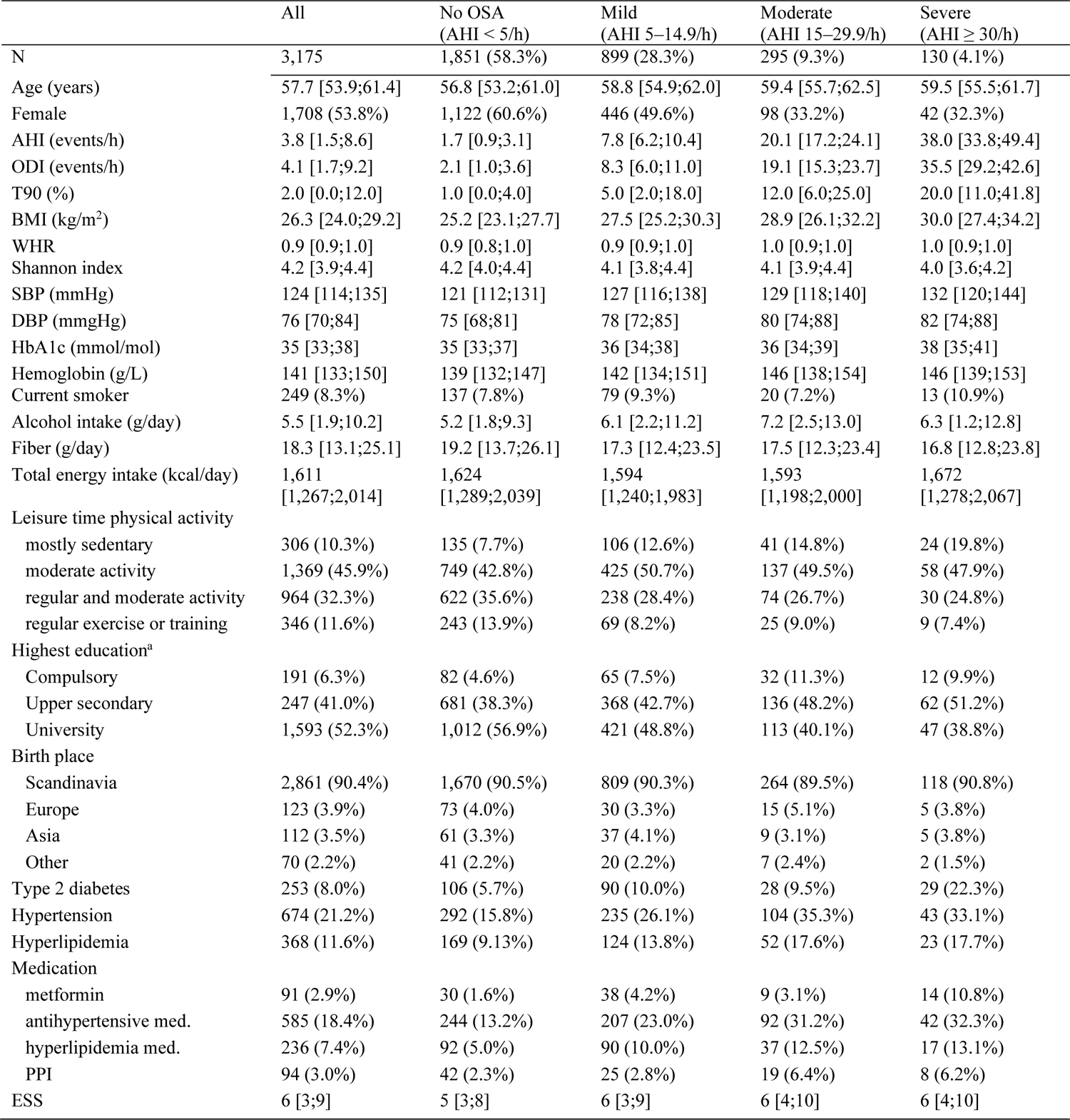
Participants’ characteristics by obstructive sleep apnea (OSA) severity groups. **caption:** Continuous variables presented as median [interquartile range] and categorical variables presented as absolute numbers (%). AHI: apnea-hypopnea index; BMI: Body mass index; DBP: diastolic blood pressure; ESS: Epworth sleepiness scale; HbA1c: glycated hemoglobin; med.: medication; ODI: oxygen desaturation index; PPI: proton-pump inhibitors; SBP: systolic blood pressure; T90: percentage of time with oxygen saturation < 90%; WHR: waist-hip-ratio. ^a^ Percentages do not add to 100% as participants with incomplete compulsory education were not included in the table.

|  | All | No OSA<br>(AHI < 5/h) | Mild<br>(AHI 5–14.9/h) | Moderate<br>(AHI 15–29.9/h) | Severe<br>(AHI ≥ 30/h) |
| --- | --- | --- | --- | --- | --- |
| N | 3,175 | 1,851 (58.3%) | 899 (28.3%) | 295 (9.3%) | 130 (4.1%) |
| Age (years) | 57.7 [53.9;61.4] | 56.8 [53.2;61.0] | 58.8 [54.9;62.0] | 59.4 [55.7;62.5] | 59.5 [55.5;61.7] |
| Female | 1,708 (53.8%) | 1,122 (60.6%) | 446 (49.6%) | 98 (33.2%) | 42 (32.3%) |
| AHI (events/h) | 3.8 [1.5;8.6] | 1.7 [0.9;3.1] | 7.8 [6.2;10.4] | 20.1 [17.2;24.1] | 38.0 [33.8;49.4] |
| ODI (events/h) | 4.1 [1.7;9.2] | 2.1 [1.0;3.6] | 8.3 [6.0;11.0] | 19.1 [15.3;23.7] | 35.5 [29.2;42.6] |
| T90 (%) | 2.0 [0.0;12.0] | 1.0 [0.0;4.0] | 5.0 [2.0;18.0] | 12.0 [6.0;25.0] | 20.0 [11.0;41.8] |
| BMI (kg/m<sup>2</sup>) | 26.3 [24.0;29.2] | 25.2 [23.1;27.7] | 27.5 [25.2;30.3] | 28.9 [26.1;32.2] | 30.0 [27.4;34.2] |
| WHR | 0.9 [0.9;1.0] | 0.9 [0.8;1.0] | 0.9 [0.9;1.0] | 1.0 [0.9;1.0] | 1.0 [0.9;1.0] |
| Shannon index | 4.2 [3.9;4.4] | 4.2 [4.0;4.4] | 4.1 [3.8;4.4] | 4.1 [3.9;4.4] | 4.0 [3.6;4.2] |
| SBP (mmHg) | 124 [114;135] | 121 [112;131] | 127 [116;138] | 129 [118;140] | 132 [120;144] |
| DBP (mmHg) | 76 [70;84] | 75 [68;81] | 78 [72;85] | 80 [74;88] | 82 [74;88] |
| HbA1c (mmol/mol) | 35 [33;38] | 35 [33;37] | 36 [34;38] | 36 [34;39] | 38 [35;41] |
| Hemoglobin (g/L) | 141 [133;150] | 139 [132;147] | 142 [134;151] | 146 [138;154] | 146 [139;153] |
| Current smoker | 249 (8.3%) | 137 (7.8%) | 79 (9.3%) | 20 (7.2%) | 13 (10.9%) |
| Alcohol intake (g/day) | 5.5 [1.9;10.2] | 5.2 [1.8;9.3] | 6.1 [2.2;11.2] | 7.2 [2.5;13.0] | 6.3 [1.2;12.8] |
| Fiber (g/day) | 18.3 [13.1;25.1] | 19.2 [13.7;26.1] | 17.3 [12.4;23.5] | 17.5 [12.3;23.4] | 16.8 [12.8;23.8] |
| Total energy intake (kcal/day) | 1,611<br>[1,267;2,014] | 1,624<br>[1,289;2,039] | 1,594<br>[1,240;1,983] | 1,593<br>[1,198;2,000] | 1,672<br>[1,278;2,067] |
| Leisure time physical activity |  |  |  |  |  |
| mostly sedentary | 306 (10.3%) | 135 (7.7%) | 106 (12.6%) | 41 (14.8%) | 24 (19.8%) |
| moderate activity | 1,369 (45.9%) | 749 (42.8%) | 425 (50.7%) | 137 (49.5%) | 58 (47.9%) |
| regular and moderate activity | 964 (32.3%) | 622 (35.6%) | 238 (28.4%) | 74 (26.7%) | 30 (24.8%) |
| regular exercise or training | 346 (11.6%) | 243 (13.9%) | 69 (8.2%) | 25 (9.0%) | 9 (7.4%) |
| Highest education<sup>a</sup> |  |  |  |  |  |
| Compulsory | 191 (6.3%) | 82 (4.6%) | 65 (7.5%) | 32 (11.3%) | 12 (9.9%) |
| Upper secondary | 247 (41.0%) | 681 (38.3%) | 368 (42.7%) | 136 (48.2%) | 62 (51.2%) |
| University | 1,593 (52.3%) | 1,012 (56.9%) | 421 (48.8%) | 113 (40.1%) | 47 (38.8%) |
| Birth place |  |  |  |  |  |
| Scandinavia | 2,861 (90.4%) | 1,670 (90.5%) | 809 (90.3%) | 264 (89.5%) | 118 (90.8%) |
| Europe | 123 (3.9%) | 73 (4.0%) | 30 (3.3%) | 15 (5.1%) | 5 (3.8%) |
| Asia | 112 (3.5%) | 61 (3.3%) | 37 (4.1%) | 9 (3.1%) | 5 (3.8%) |
| Other | 70 (2.2%) | 41 (2.2%) | 20 (2.2%) | 7 (2.4%) | 2 (1.5%) |
| Type 2 diabetes | 253 (8.0%) | 106 (5.7%) | 90 (10.0%) | 28 (9.5%) | 29 (22.3%) |
| Hypertension | 674 (21.2%) | 292 (15.8%) | 235 (26.1%) | 104 (35.3%) | 43 (33.1%) |
| Hyperlipidemia | 368 (11.6%) | 169 (9.13%) | 124 (13.8%) | 52 (17.6%) | 23 (17.7%) |
| Medication |  |  |  |  |  |
| metformin | 91 (2.9%) | 30 (1.6%) | 38 (4.2%) | 9 (3.1%) | 14 (10.8%) |
| antihypertensive med. | 585 (18.4%) | 244 (13.2%) | 207 (23.0%) | 92 (31.2%) | 42 (32.3%) |
| hyperlipidemia med. | 236 (7.4%) | 92 (5.0%) | 90 (10.0%) | 37 (12.5%) | 17 (13.1%) |
| PPI | 94 (3.0%) | 42 (2.3%) | 25 (2.8%) | 19 (6.4%) | 8 (6.2%) |
| ESS | 6 [3;9] | 5 [3;8] | 6 [3;9] | 6 [4;10] | 6 [4;10] |

### OSA is associated with lower gut microbiota richness and evenness

To investigate whether OSA was associated with the richness and evenness of the gut microbiota, we performed partial Spearman’s correlation analyses of the three OSA parameters a) AHI, b) T90, and c) ODI with alpha diversity measured as the Shannon index^22^. We used a hypothetical causal diagram (Fig. S2) and the d-separation criterion^23^ to identify the minimal set of confounders for adjustment in our main model. Thus, covariates in the main model consisted of age, sex, smoking, alcohol intake, and BMI, as well as the DNA extraction plate to account for variation between batches. We found that AHI, T90, and ODI were inversely associated with the Shannon index (AHI: ρ = −0.058, p-value = 0.002; T90: ρ = −0.043, p-value = 0.013, ODI: ρ = − 0.065, p-value = 1.75×10^-4^, Table S3). To better account for the influence of diet and other confounders, we also constructed an extended model with additional adjustments for calculated fiber intake and total energy intake from a food frequency questionnaire, self-reported leisure time physical activity, education level, country of birth, and season of assessment. In this extended model, the associations were somewhat attenuated (AHI: ρ = −0.047, p-value = 0.013; T90: ρ = −0.038, p-value = 0.034, ODI: ρ = −0.055, p-value = 0.002). Taken together, these results indicate a decreased gut microbiota richness and evenness in OSA.

### Gut microbiota composition differs by OSA severity independently of BMI

To determine whether the OSA severity was associated with the overall gut microbiota composition, we analyzed the beta diversity measured as Bray-Curtis dissimilarity in relation to groups of AHI, T90, and ODI. Starting with AHI, we grouped the participants (n=3,175) with valid AHI data according to previously established and clinically used cut-off values^24^ (no OSA: AHI < 5, n = 1,851; mild: AHI 5–14.9, n = 899; moderate: AHI 15–29.9, n = 295; severe: AHI ≥ 30, n = 130). To graphically represent the relationship between the groups, we conducted a principal coordinate analysis on the Bray-Curtis dissimilarity matrix (Fig. 1a). We observed a separation of the groups along the first axis in order of severity. This separation was supported by the results of a permutational analysis of variance (PERMANOVA) adjusted for the main model covariates (R^2^ = 0.5% and p-value = 0.0001). In pairwise comparisons (Table S4), we found some evidence supporting a difference for all pairwise comparisons (nominal p-value < 0.05), except between the groups mild and severe (p-value = 0.26), probably due to higher dispersion and lower power in this comparison. Taken together, these results suggest that the groups of OSA severity based on AHI have different gut microbiota compositions.

**Figure 1.**
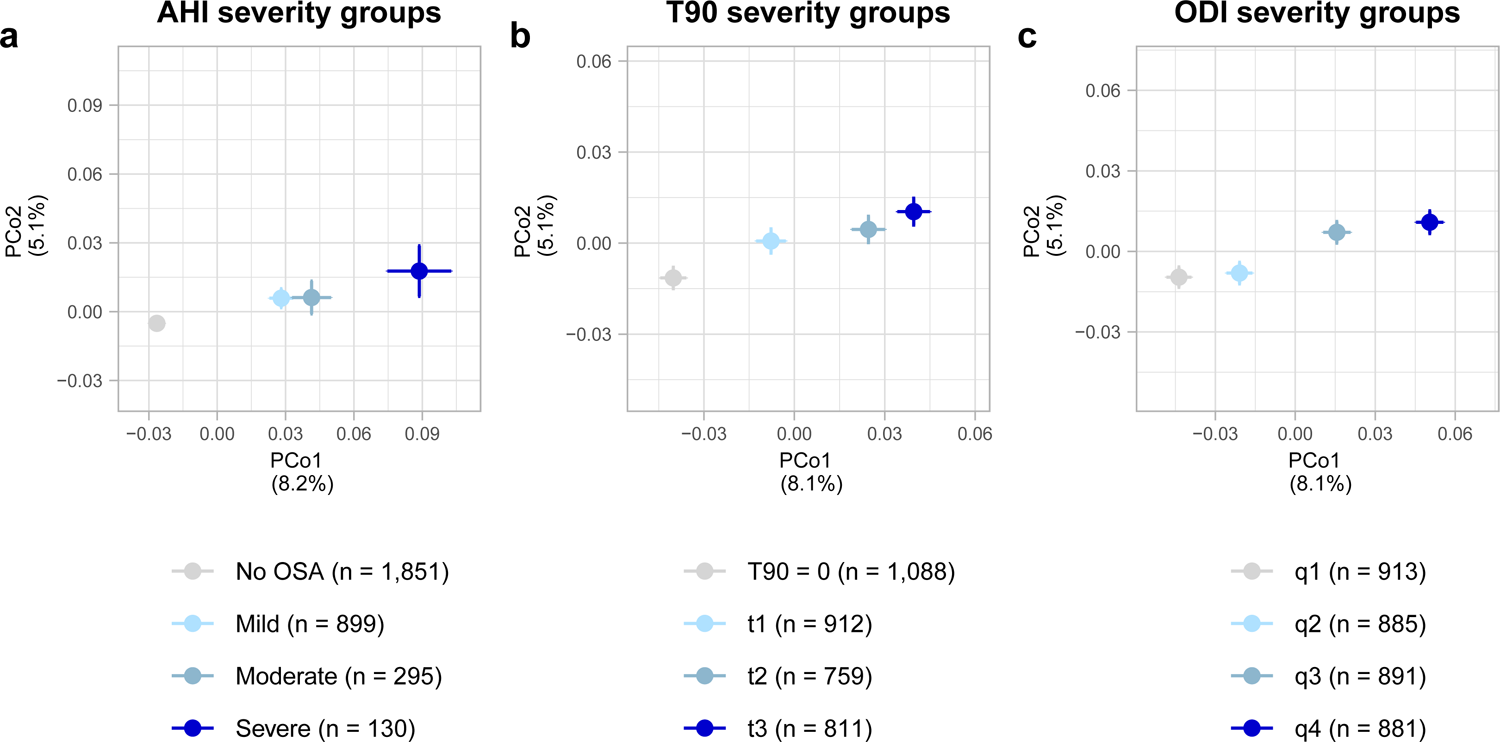
Principal coordinate analysis showing progressive differences in the overall gut microbiota composition, measured with Bray-Curtis dissimilarity, across groups of obstructive sleep apnea (OSA) severity. Closed circles represent the mean axis value per group and bars represent the standard errors. The percentages on the axes labels represent the variance explained by each axis. **a**. AHI severity groups. No OSA: AHI < 5; Mild: AHI 5–14.9; Moderate: AHI 15– 29.9; Severe: AHI ≥ 30. **b.** T90 severity groups were defined as one category including participants with T90 = 0, and the remaining participants divided by tertiles (t1: T90 = 1–3; t2: T90 = 4–14; and t3: T90 ≥ 15). **c.** ODI severity groups were defined by quartiles of ODI (q1: ODI = 0–1.8; q2: ODI = 1.9–4.3; q3: ODI = 4.4–9.4; and q4: ODI ≥ 9.5). AHI: apnea-hypopnea index; ODI: oxygen desaturation index; PCo: principal coordinate; T90: percentage of time with oxygen saturation < 90%.

To assess whether the gut microbiota composition also differed among participants depending on their hypoxia parameters, we divided the participants with available T90 and ODI data (n=3,570) into four groups. Because there were no well-established cut-offs, we aim to create groups of similar sizes. Due to the high proportion of participants with T90 = 0, grouping by quartiles was not possible for this variable. Therefore, we performed the following division for T90: one group of participants with T90 = 0 (n=1,088), and the remaining participants divided into three groups of similar size in order of ascending T90 values: t1 = 912 individuals (T90: 1–3%), t2 = 759 (T90: 4–14%), and t3 = 811 (T90: 15–100%). For ODI, we divided the participants by quartiles: q1 = 913 individuals (ODI: 0–1.8), q2 = 885 (ODI: 1.9–4.3), q3 = 891 (ODI: 4.4–9.4), and q4 = 881 (ODI: 9.5–93). Either based on T90 or ODI, the groups separated along the first axis in order of severity in the principal coordinate analysis (Fig. 1b and 1c). We confirmed the separations using PERMANOVA (T90 groups: R^2^ = 0.2% and p-value = 0.013. ODI groups: R^2^ = 0.3% and a p-value = 0.002). In the pairwise comparisons of T90 groups (Table S4), we found that the group with T90 = 0 differed from the t2 and t3 groups (p-values = 0.001 and 0.006, respectively). In the pairwise comparisons of ODI groups (Table S4), we found that the q1 group differed from the q3 and q4 groups (p-values = 0.014 and 0.001, respectively). We also found a difference between the q2 and q4 groups (p-value = 0.005).

We additionally investigated differences in the overall gut microbiota between the groups after adjusting for the extended model covariates. For all grouping strategies; i.e., AHI-, T90- or ODI-based, we confirmed the differences in gut microbiota composition across groups (AHI groups: R^2^ = 0.5%, p-value = 0.0001; T90 groups: R^2^ = 0.2%, p-value = 0.028; ODI groups: R^2^ = 0.2%, p-value = 0.007).

Altogether, these results point to progressive differences in the overall gut microbiota composition – as assessed with Bray-Curtis dissimilarity – across groups of OSA ordered by severity. The difference between groups was clearer when using the pre-established AHI cut-offs rather than the similarly sized groups based on T90 and ODI. We could still detect differences in the gut microbiota composition between groups of T90 or ODI even after accounting for several confounders.

### OSA-related hypoxia is associated with the relative abundance of specific gut microbiota species independently of BMI, dietary fiber intake, and common medications

To study how the OSA parameters AHI, T90, and ODI were associated with the relative abundance of gut microbiota species, we performed a series of partial Spearman’s correlations with each OSA parameter and each species. Given that BMI may influence or be influenced by the abundance of gut microbiota species^25^ and that obesity is an important cause of OSA^3^, BMI could either be considered a confounder or a source of reverse causation in the association of OSA with microbiota. Therefore, we divided the main model into two: one model not including, and one model including BMI. The model not including BMI was then used as a screening step to select the species that would be taken into the subsequent models. We defined significance using p-values adjusted for multiple comparisons, referred to here as q-values, with a false discovery rate set at 5%.

Without adjustment for BMI, we found that AHI was associated with the relative abundances of 566 species, T90 with 631 species, and ODI with 692 species (Fig. 2a and Table S5). Next, the species that were associated with at least one of the OSA parameters in this step were analyzed with the main model including BMI. In these analyses, AHI was associated with 101 species, T90 with 141, and ODI with 241 (Table S6). When assessing the overlap of these associations, we found that the three OSA parameters were jointly associated with the relative abundances of 53 species. The AHI and ODI parameters were jointly associated with another 42 species, ODI and T90 were jointly associated with another 46 species, and AHI and T90 were jointly associated with one other species (Fig. 2b).

**Figure 2.**
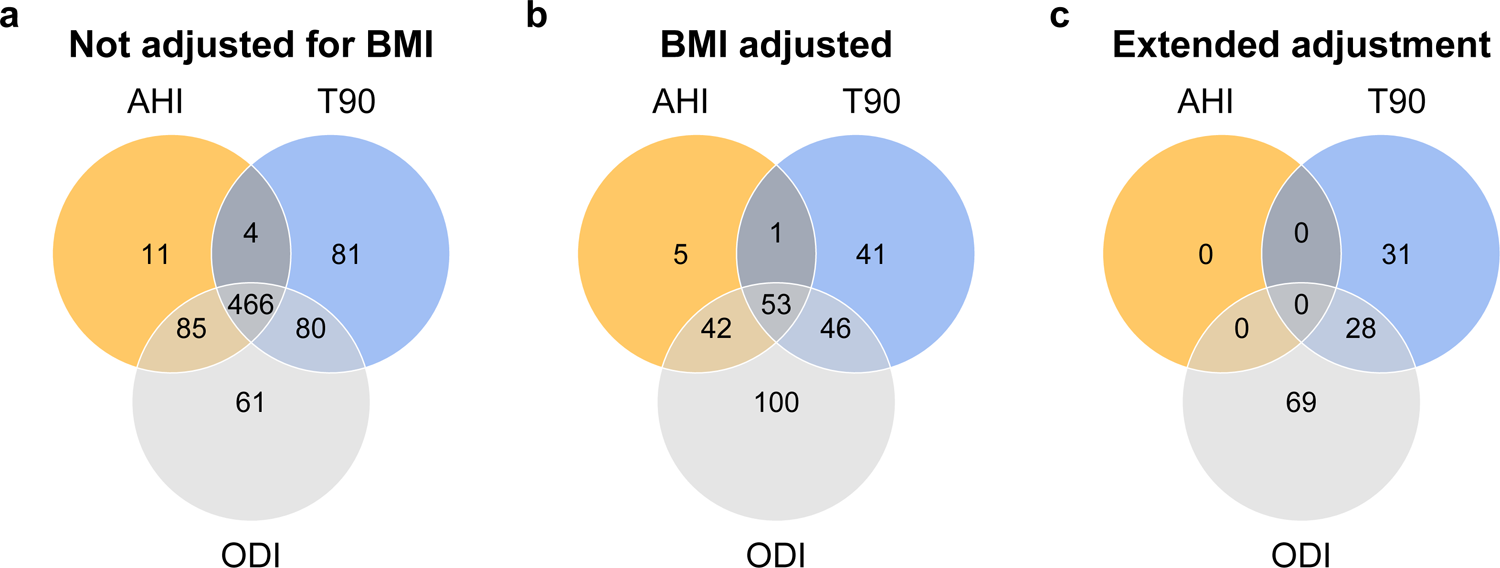
Number of microbiota species associated with AHI, T90, and/or ODI. Associations investigated using partial Spearman’s correlation after removing participants with missing data on covariates. Adjustment for multiple comparisons using the Benjamini-Hochberg method with a 5% false discovery rate. **a.** Results from the main model (i.e., adjustment for age, sex, smoking, alcohol intake, and DNA extraction plate) not including adjustment for body mass index (BMI) and **b.** including adjustment for BMI. For AHI, n = 3004. For T90 and ODI, n = 3,364. **c.** Results from the extended model (i.e., further adjustment for fiber intake, total energy intake, leisure time physical activity, education, country of birth, and season). For AHI, n = 2,909. For T90 and ODI, n = 3,249. AHI: apnea-hypopnea index; ODI: oxygen desaturation index; T90: percentage of time with oxygen saturation < 90%.

To account for other potential confounders, the species associated with at least one of the OSA parameters in the main model not including BMI were then analyzed with the extended model. In these analyses, AHI was no longer associated with any species (Fig. 2c and Table S7). Nevertheless, T90 was associated with 59 species, of which 17 were positive, and ODI with 97 species, of which 17 were positive. The parameters T90 and ODI were jointly positively associated with six species, including the species *Blautia obeum* (internal identifier: HG3A.0001), *Ruminococcus gnavus*, *Coprococcus comes*, and the recently isolated *Mediterraneibacter glycyrrhizinilyticus*^26^. The T90 and ODI parameters were jointly negatively associated with 22 unidentified species, of which 16 belonged to the order *Eubacteriales*. Based on the correlation coefficients, the strongest positive associations with ODI were *Fusicatenibacter saccharivorans* (ρ = 0.069, q-value = 0.008), and an unidentified species of the family *Lachnospiraceae* (HG3A.0018, ρ = 0.069, q-value = 0.008). Among the 17 positive associations with ODI, 10 species belonged to the family *Lachnospiraceae*. The strongest negative association of ODI was with an unidentified *Eubacteriales* sp. (HG3A.0069, ρ = −0.074, q-value = 0.006). The strongest positive association of T90 was with *Dorea formicigenerans* (ρ = 0.086, q-value = 0.001), followed by *B. obeum* (HG3A.0001, ρ = 0.079, q-value = 0.003). The strongest negative association was with *Eubacteriales* sp. (HG3A.0703, ρ = −0.073, q-value = 0.009).

One possible explanation for the null findings for AHI was that fewer participants had valid AHI data compared with the number of participants with valid T90 and ODI data. To ascertain that the null findings were not due to lower power, we conducted a secondary analysis where we used multiple imputations to impute missing AHI values for the 340 participants who had valid T90 or ODI values, but not valid AHI values. Even in this secondary analysis, we did not observe any associations between AHI and species after adjustment for the extended model covariates (Table S8).

Next, we performed a series of sensitivity analyses with the 128 species associated with T90 and/or ODI in the extended model to investigate how medication use or presence of lung disease affected our findings (Table S9). Firstly, we added to the extended model the covariates of metformin use, and proton pump inhibitor (PPI) use, based on measurable metabolomic plasma levels of these medications, as well as self-reported medication use for hypertension and/or hyperlipidemia. All 128 species remained associated with T90 and/or ODI after the additional adjustment (q-value < 0.05). After excluding the 367 participants who had used any antibiotic during the six months before sampling, all associations were retained, except for the association between ODI and *Eubacteriales* sp. (HG3A.0691). The result also did not change after excluding the 29 participants with self-reported diagnosis of chronic obstructive pulmonary disease (COPD), chronic bronchitis, or pulmonary emphysema. Altogether, this series of analyses highlighted the robustness of the associations between the hypoxia parameters T90 and ODI and the identified species. In summary, we found that the OSA-related hypoxia parameters were robustly associated with 127 gut microbiota species.

### The T90 and ODI associations with gut microbiota species were not modified by hemoglobin levels

Exposure to hypoxia increases erythropoiesis and results in increased hemoglobin levels^27^. In turn, hemoglobin level affects the oxygen delivery to tissues^28^. To assess the effect modification by hemoglobin levels on the T90 and ODI associations with the gut microbiota species, we conducted Spearman’s correlation analyses adjusted for the extended model covariates stratified by the sex-specific median hemoglobin value. These analyses included the 128 species that we identified as being associated with the T90 and/or ODI parameters (Table S10). In the association between ODI and *Eubacteriales* sp. (HG3A.1026), we detected a difference in the correlation estimates between participants with high or low hemoglobin status (ρ_low_ = 0.008, p-value_low_ = 0.75 and and ρ_high_ = −0.12, p-value_high_ =1.8×10^-6^, heterogeneity q-value = 0.02).

Differences were also observed for other species, as in the association between ODI and *Clostridia* sp. (HG3A.0140, ρ_low_ = −0.12, p-value_low_ = 4.0×10^-6^ and ρ_high_ = −0.02, p-value_high_ = 0.36), but when we formally tested for these differences, we could not confirm them (heterogeneity q-value > 0.05). Therefore, we could not ascertain whether hemoglobin levels acted as an effect modifier on the associations between OSA-related hypoxia parameters and the gut microbiota species abundance.

### T90-associated gut microbiota showed enriched putative metabolic pathways

To characterize the putative metabolic profile of the species associated with OSA, we performed enrichment analyses to identify overrepresented metabolic pathways among the AHI, T90, or ODI associations with gut microbiota species (extended model). Metabolic pathways were defined using the gut metabolic modules (GMM)^29^. We found no metabolic pathways enriched in the AHI or ODI associations (Table S9). The positive associations between T90 and the gut microbiota species were enriched for nine metabolic pathways (Fig. 3a), including threonine degradation I and II (q-value = 9.3×10^-4^ and 0.02, respectively), and propionate production II (q-value = 0.02). The latter is comprised of the enzyme propionate CoA-transferase, which produces propionate from lactate^30^. The pathway threonine degradation I consists of a series of enzymatic steps that metabolize L-threonine into propionate. One of the these steps is carried out by the activated pyruvate-formate lyase, which is only activated under anaerobic conditions^31^. The T90-species associations were also enriched for lysine degradation II (q-value = 0.004), serine degradation (q-value = 0.009), and the pathways of carbohydrate degradation: galacturonate degradation I (q-value < 0.001, and ribose degradation (q-value = 0.047).

**Figure 3.**
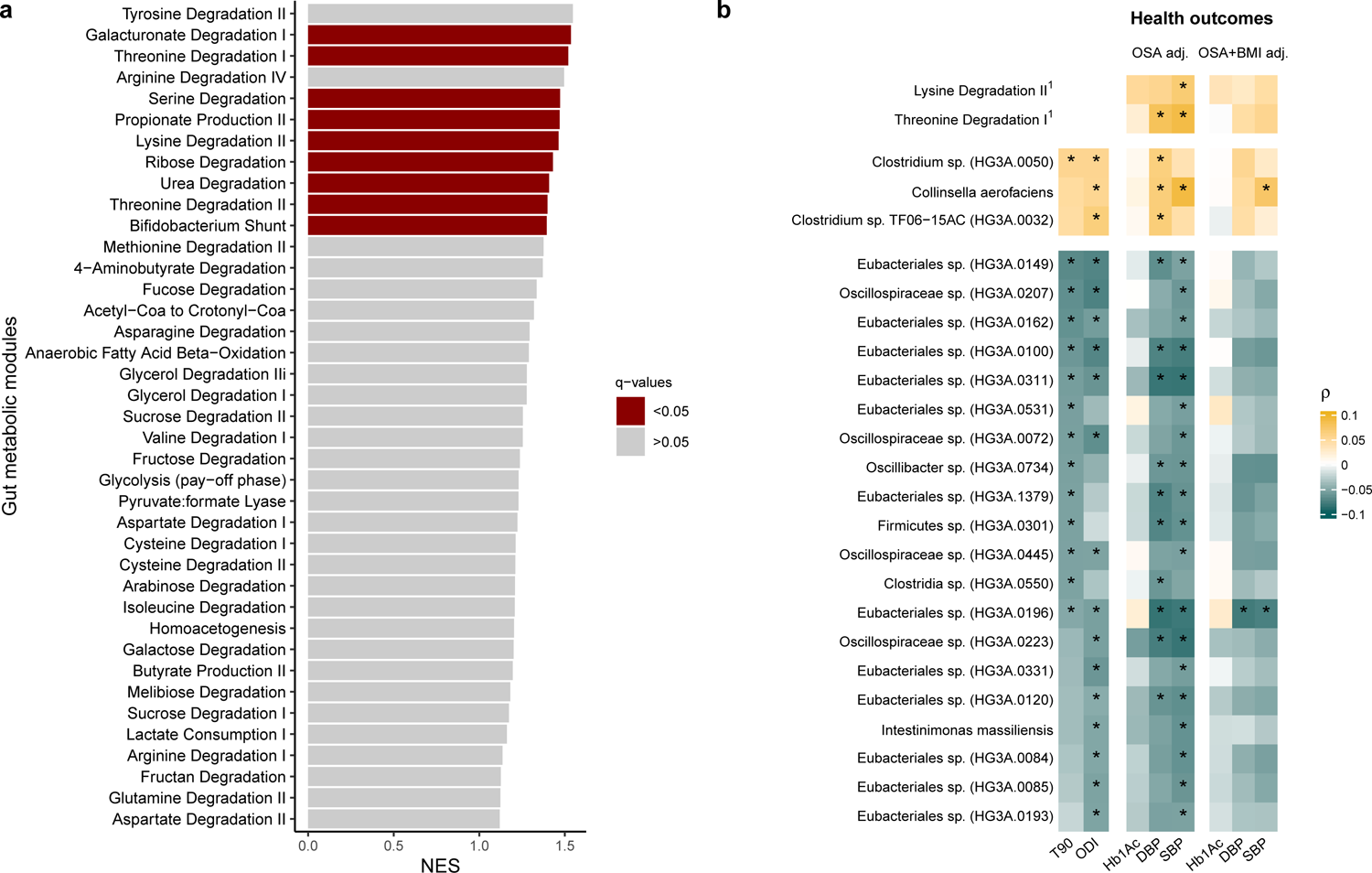
**a.** Enrichment for gut metabolic modules (GMM) among the positive associations between T90 and gut microbiota species. The pathway enrichment analysis was conducted on the ranked p-values obtained from Spearman’s correlations adjusted for age, sex, alcohol intake, smoking, body mass index, fiber intake, total energy intake, leisure time physical activity, education, country of birth, season, and DNA extraction plate. Multiple comparisons were adjusted for using the Benjamini-Hochberg method. q-values: adjusted p-values considering a 5% false discovery rate **b**. Heatmap showing the species associated with T90 or ODI and the T90-enriched GMM(^1^) that were also associated with at least one of the health outcomes: systolic blood pressure (SBP), diastolic blood pressure (DBP), and glycated hemoglobin (Hb1Ac). OSA adj: Spearman’s correlation adjusted for age, sex, alcohol intake, smoking, fiber intake, total energy intake, leisure time physical activity, country of birth, apnea-hypopnea index (AHI), oxygen desaturation index, T90, and DNA extraction plate. OSA+BMI adj: additional adjustment for body mass index (BMI). Associations with an asterisk (*) were identified considering a 5% false discovery rate. NES: normalized enrichment score; OSA: obstructive sleep apnea; T90: percentage of time with oxygen saturation < 90%.

Our results on the enrichment of microbial metabolic pathways suggest that the duration of hypoxia during sleep, as reflected by the T90 parameter, may favor gut microbiota species with specific metabolic repertoires. Noteworthy, we found that T90 was associated with the production of propionate from lactate, a biomarker of hypoxia. On the other hand, we could not find evidence that AHI or ODI were associated with metabolic features of the gut microbiota.

### Species positively associated with T90 and/or ODI have different metabolomic fingerprints than those negatively associated

To characterize the metabolomic fingerprint of the 128 gut microbiota species associated with T90 and/or ODI in the extended model, we mined data from the online GUTSY Atlas (https://gutsyatlas.serve.scilifelab.se/), which has investigated the associations between the human gut microbiota and the plasma metabolome in a cohort that also included participants from the current study^32^. From the GUTSY Atlas, we retrieved data on the enrichment analyses for metabolite groups in the associations between the species and the plasma metabolites. The analyses were conducted stratified by the direction of the associations.

In the heatmaps of enriched metabolite groups for every species, we observed a different pattern of enrichment for the 100 species that had a negative association with T90 and/or ODI values compared with the 28 species that had a positive association with these parameters. For instance, while several positively-associated species were positively associated with secondary bile acids (Fig. 4b), the negatively-associated species were negatively associated with these metabolites (Fig. 4a). A similar pattern was also observed for dihydrosphingomyelins and phosphatidylcholine metabolites. In addition, for 19 species that were negatively associated with T90/ODI, we found enrichment for vitamin A metabolites; whereas four species in positive association with T90/ODI had a negative association with vitamin A metabolites.

**Figure 4.**
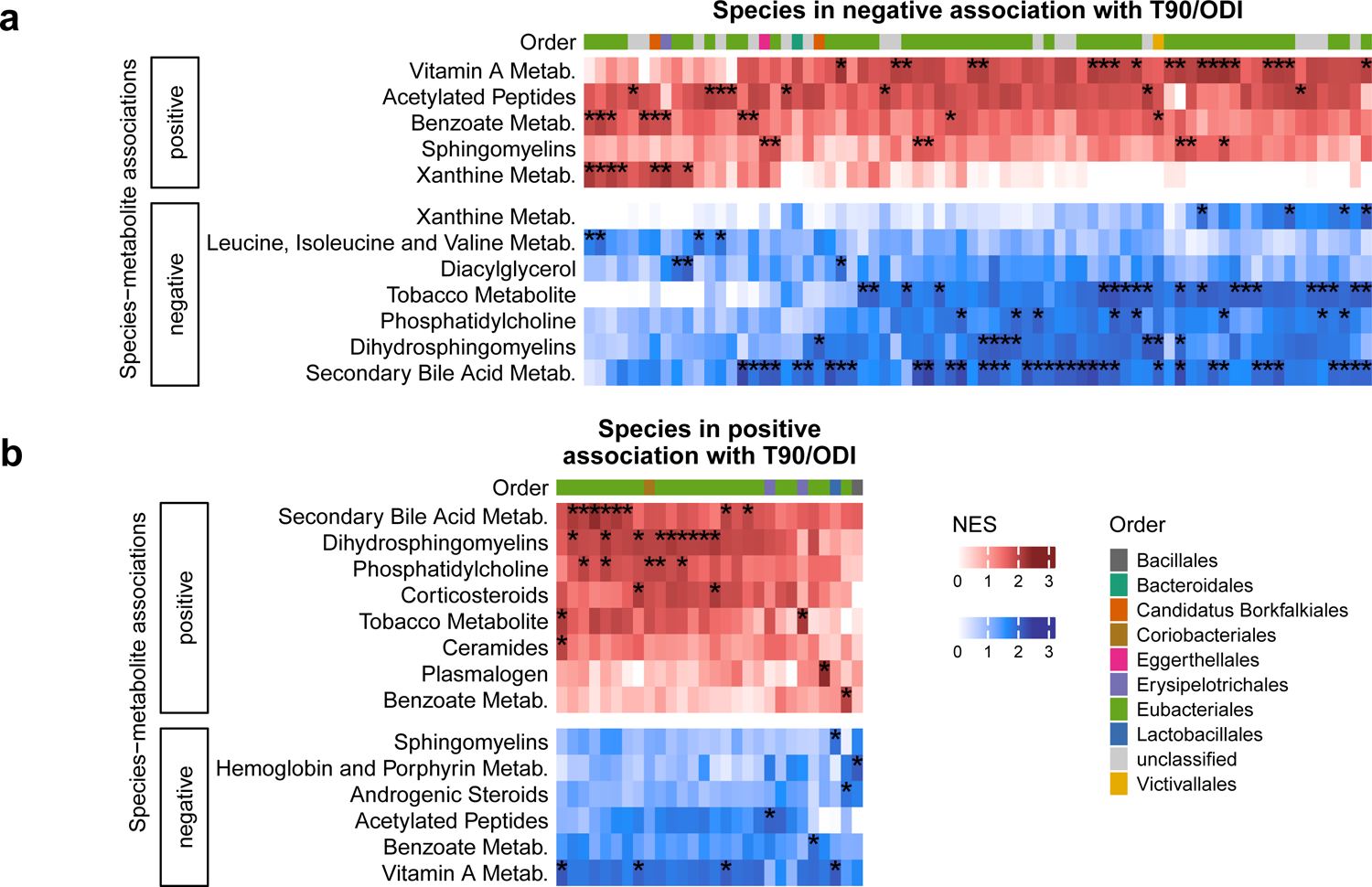
Relationship of the T90- and/or ODI-associated species with the plasma metabolome. T90 and ODI were **a.** negatively associated with 100 species and **b.** positively associated with 28 species. The heatmaps show the enriched metabolites groups in the associations between species and metabolites stratified by the direction of the associations. Enrichment results retrieved from the GUTSY Atlas (https://gutsyatlas.serve.scilifelab.se/)32. Multiple comparisons were adjusted for using the Benjamini-Hochberg method. Associations with an asterisk (*) were identified considering a false discovery rate of 5%. NES: normalized enrichment score; ODI: oxygen desaturation index; T90: percentage of time with oxygen saturation < 90%.

Lastly, we found evidence that the species that were negatively associated with the hypoxia parameters T90 or ODI were also negatively associated with tobacco metabolites. Although these species were identified in analyses adjusted for smoking status, it is possible that these species may also have a reduced abundance in smokers, hence the negative association with tobacco metabolites.

Overall, our results indicated that the associations between the species and the plasma metabolites had opposite directions depending if the species abundance was expected to increase or decrease with hypoxia. The main differences in the metabolomic fingerprints involved the pattern of association with vitamin A metabolites, dihydrosphingomyelins, secondary bile acids, phosphatidylcholine, and tobacco metabolites.

### Microbiota species associated with OSA-related hypoxia and were also associated with blood pressure

Previous studies have found that OSA is associated with increased risk for insulin resistance^33^ and incident blood hypertension^4^. Furthermore, fecal microbiota transplantation studies in mice have suggested that alterations in the gut microbiota induced by OSA might contribute to the detrimental health effects of OSA^15, 16^. Therefore, we investigated whether the 128 T90/ODI-associated species and the nine T90-enriched metabolic pathways were also associated with blood pressure measurements (n = 2,335) and glycated hemoglobin (Hb1Ac, n = 2,786). We excluded from the association analyses with blood pressure measurements the participants who self-reported medication for hypertension. Likewise, we excluded from the analyses with HbA1c the participants who self-reported medication for diabetes. Based on a directed acyclic graph (Fig. S3), the associations were adjusted for age, sex, alcohol intake, smoking, fiber intake, total energy intake, leisure time physical activity, country of birth, ODI, AHI, T90, and DNA extraction plate. Among the species negatively associated with T90 or ODI, 19 were negatively associated with systolic blood pressure (Fig. 3b), including *Intestinimonas massiliensis* (ρ = − 0.07, q-value = 0.02), and nine were negatively associated with diastolic blood pressure (Table S12). Among the positive associations, the species *Collinsella aerofaciens*, and the microbial pathway threonine degradation I were positively associated with systolic and diastolic blood pressure. None of the species or microbiota functions investigated were associated with Hb1Ac.

In the analyses additionally adjusted for BMI, *C. aerofaciens* continued to associate with systolic blood pressure (ρ = 0.073, q-value = 0.034) and *Eubacteriales* sp. (HG3A.0196) continued to negatively associate with systolic (ρ = −0.077, q-value = 0.032) and diastolic blood pressure (ρ = −0.081, q-value = 0.015). Altogether, we could observe that specific species and metabolic pathways previously identified in association with T90 or ODI were also associated with blood pressure measurements in the same direction. Two species continued to be associated with blood pressure measurements even after adjustment for BMI.

## Discussion

Here we presented the most comprehensive population-based study to date investigating the relationship of OSA with the human gut microbiota. We found evidence that OSA, especially OSA-related hypoxia, was associated with the composition and functional potential of the human gut microbiota. The OSA hypoxia parameters, namely T90 and ODI, were associated with the abundance of specific species after extensive adjustment for potential confounders such as treatment for diabetes, hypertension, hyperlipidemia, and gastritis and gastroesophageal reflux. We further noted enrichment for specific microbial metabolic pathways shared by bacteria positively associated with the duration of hypoxia during sleep.

Out of the 128 species associated with T90 and/or ODI, 28 were associated with both parameters. Among the six positive associations, four were annotated to the species level, namely *B. obeum*, *C. comes*, *R. gnavus*, and *M. glycyrrhizinilyticus*. A higher relative abundance of *B. obeum*, previously named *Ruminococcus obeum*, has been observed in individuals with insulin resistance^34^. Nevertheless, we did not find an association between *B. obeum* and HbA1c levels, a marker of glycemic control, in the present study. *C. comes* is one of the gut microbiota species found to decrease in abundance post bariatric surgery^35^, a procedure known to improve many conditions related to obesity, including OSA^36^. *R. gnavus*, a mucin-degrading microbe, has been positively associated with incident type 2 diabetes in a large Finnish population cohort^37^.

These four species all belong to the family *Lachnospiraceae*. An increased abundance of the *Lachnospiraceae* family has been observed in mice subjected to intermittent hypoxia^12^. Also belonging to the same family are *D. formicigenerans* and *Anaerobutyricum hallii*, which were positively associated with T90; and *Roseburia inulinivorans*, *F. saccharivorans*, *Ruminococcus torques* and *Blautia massiliensis*, which were positively associated with ODI. However, a mechanistic explanation connecting the *Lachnospiraceae* family to host hypoxia is lacking and warrants further investigation.

In this study, ODI was positively associated with *C. aerofaciens*, which was in turn positively associated with systolic blood pressure independent of BMI and OSA. *C. aerofaciens* is an obligate anaerobe abundant in the human gut^38^, with higher abundance in overweight and obese individuals^39^. Higher abundance of *C. aerofaciens* has also been observed in individuals with type 1 pulmonary hypertension^40^, a condition associated with lower oxygen saturation^41^. Longitudinal and experimental studies are necessary to understand whether the gut microbiota species identified in this study may have a role connecting OSA to blood pressure.

The OSA hypoxia parameters T90 and ODI were associated with the abundance of specific species after adjustment for the extended model covariates, while AHI was not. Our results suggest that OSA-related hypoxia might be of greater importance when it comes to associations with the gut microbiota than the apnea and hypopnea events themselves. Increasing attention has been given to OSA-related hypoxia, as recent studies have shown that OSA-related hypoxia, unlike AHI, predicted increased cardiovascular mortality^5^.

Evidence in the literature suggests that intermittent host hypoxia directly affects the gut microbiota. In the study by Moreno-Indias et al., intermittent hypoxia applied during the mice’s rest phase resulted in enrichment for gut obligate anaerobes^12^. Similarly, a reduction of anaerobes was observed in the gut microbiota of rats after hyperbaric treatment^42^. In the Moreno-Indias et al. study, the authors demonstrated that intermittent hypoxia produced oscillations in the oxygen concentration inside the intestinal lumen close to the epithelium, thus providing a physiological rationale for how the oxygenation level of the host could impact the gut microbiota environment^12^.

The identified associations could be due to effects of OSA on sleep fragmentation as both acute and chronic sleep fragmentation have been demonstrated to affect the gut microbiota composition in rodents^43^. Nevertheless, sleep fragmentation alone is unlikely to explain why OSA hypoxia parameters associated with the abundance of specific species while AHI did not. Daytime sleepiness may be a result of sleep fragmentation; however, the Epworth sleepiness scale score, a measurement of daytime sleepiness, was not different between the groups of OSA severity based on T90 (Table S1). To better disentangle the effects of sleep fragmentation due to OSA from the effects of nocturnal hypoxia, future studies would benefit from concomitant electroencephalogram monitoring, polysomnography assessment and actigraphy.

One possible indirect mechanism through which OSA might affect the microbiota is through alterations in the host metabolism, such as accumulation of lactate^44^. Experiments in mice using labeled lactate showed that circulating lactate can cross the gut barrier and enter the intestinal lumen^45^. Higher plasma concentration of lactic acid has benn described in OSA patients^46^. Additionally, T90 was positively associated with venous concentration of lactate in treatment-naïve OSA patients^44^. High circulating levels of lactate is also present in athletes postexercise; although, in this case, lactate is produced in the absence of hypoxia^47^. A study comparing the gut microbiota in athletes before and after exercise found an increase in microbial genes involved in the conversion of lactated into propionate after exercise^45^. The authors of the latter study hypothesized that the host lactate entering the intestinal lumen could favor bacterial species that use lactate as a carbon source. This hypothesis corresponds with our findings; the pathway of propionate production from lactate was enriched in positive T90 associations with gut microbiota species. In summary, supported by the existing literature, our study suggests an association between the hypoxia caused by OSA, plasma lactate, and the gut microbiota metabolic profile.

The strengths of our study include the large sample size, temporal proximity between OSA assessment and fecal microbiota sampling, the extensive adjustment for confounders, and the use of an objective assessment for OSA instead of self-reported diagnosis or a convenience sample of OSA patients. Our investigation on a random sample from the population provides a more generalizable picture of the association between OSA and the gut microbiota than studies comparing controls to patients with a clinical diagnosis of OSA. The combination of a large sample size with shotgun metagenomic sequencing allowed us to conduct a comprehensive investigation at the species level and also of the microbial metabolic profile.

Nevertheless, there are limitations that need to be considered when interpreting our results. Because of the cross-sectional design, we were not able to assess causality. Despite the extensive adjustment, we cannot rule out residual confounding. Moreover, even if the animal studies indicate a causal effect of OSA on the gut microbiota, an effect in opposite direction is also plausible. For example, certain gut microbiota species may promote weight gain that may consequently lead to OSA^48^. In this case, BMI would open a pathway for reverse causation from the gut microbiota to the development of OSA. The gut microbiota was examined using fecal shotgun metagenomics followed by detection of species based on the co-abundance of genes.

Although valid and well-described^49^, this approach leads to the detection of some incompletely annotated species, which have not yet been isolated and characterized. Alternatively to fecal samples, mucosal biopsies could have provided information on the gut microbiota that resides more closely to the host^50^, thus more likely to be affected by OSA. Our results may not extend to populations in other countries due to the close connection between geographical location and the gut microbiota^51^. The ApneaLink Air® two-channel device used to assess sleep apnea does not distinguish obstructive from central apneas. However, obstructive sleep apneas are more common in the general population^52^. An assessment for OSA based on a single night may result in a certain degree of exposure misclassification^53^, which could affect the precision, but would not bias our estimates. The T90 parameter is not able to differentiate hypoxia caused by OSA from nocturnal hypoxia of other etiologies. Finally, we excluded participants that used CPAP, a common treatment for OSA. Thus, future studies should investigate whether the herein observed OSA-gut microbiota associations are affected by OSA therapy, including but not limited to CPAP.

Animal studies have found that comorbidities present in mouse models of OSA can be transferred to control mice through fecal microbial transplant^15, 17^. Furthermore, observational studies have implicated the human gut microbiota in the development of metabolic and cardiovascular disorders, such as hypertension^54^ and insulin resistance^55^. Given the results from animal and observational studies, it is possible that alterations in the human gut microbiota caused by OSA could at least in part contribute to the increased risk of cardiovascular disease seen in OSA patients. Therefore, the associations that we found between OSA and the gut microbiota can be informative for future research aiming to identify gut microbiota mechanisms that connect OSA to health outcomes.

In conclusion, we present the largest study to date that has investigated the association between OSA and the human gut microbiota. We found that the objective parameters of OSA-related hypoxia, namely T90 and ODI, were independently associated with 59 and 97 gut microbiota species, respectively. In addition, we found that the gut microbiota associated with T90 was enriched for nine metabolic pathways, including the pathway for production of propionate from lactate. Our findings provide novel insights into the relationship between OSA and the gut microbiota. Future experimental studies are necessary to validate whether the identified microbial species may represent potential therapeutic targets to prevent or treat comorbidities of OSA.

## Methods

### Study population

From 2013 to 2018, a total of 30,154 women and men aged 50–64 were enrolled in the SCAPIS study. Participants were randomly invited from the general population across six regions in Sweden^56^. In the Uppsala region, 4,839 participants had data on fecal shotgun metagenomics and 4,183 were assessed for OSA. Combined, 4,045 participants had both OSA data and metagenomic data. We excluded the 59 participants who reported use of CPAP. At least four hours of air flow and oxygen saturation recordings were required to compute a valid AHI value and at least four hours of oxygen saturation recording was required to compute valid T90 and ODI values. Therefore, data on AHI were available for 3,175 participants and data on T90 and ODI were available for 3,570 participants (Fig. S1).

SCAPIS study and the present study were approved by the Swedish Ethical Review Authority (DNR 2018-315 B and amendment 2020-06597, and DNR 2010-228-31M, respectively). All participants provided informed consent.

### OSA assessment

Assessment for OSA was conducted using the ApneaLink Air® (ResMed, CA, USA)^57^. After instructions, participants took the ApneaLink Air® device home for recording of nasal air flow and oxygen saturation during one night’s sleep. Apnea was defined as a reduction of breathing flow ≥80% for at least 10 seconds. Hypopnea was defined as a period of at least 10 seconds with a decrease in the baseline air flow of 30–80% combined with a decrease ≥4% in oxygen saturation. From the OSA assessment, the variables AHI, T90, and ODI were chosen for the current study as they are clinically relevant measures and commonly used in previous research. The AHI was calculated as the mean number of apneas and hypopneas events per hour of total recording time. Using clinically used cuff-off values, AHI severity groups were defined as no OSA for AHI < 5, mild for AHI 5–14.9, moderate for AHI 15–29.9, and severe for AHI ≥ 30^24^. The T90 variable was computed by adding the time spent with an oxygen saturation < 90% and dividing by the total recording time. For the grouping based on T90, the first group was composed of participants with a T90 = 0. The remaining participants were grouped by tertiles. It was not possible to group the participants based on quartiles because > 25% of the participants had a T90 = 0. ODI was calculated as the mean number of desaturation events per hour during the total recording time. A desaturation event was defined as a decrease from baseline ≥ 4% in oxygen saturation. For the grouping based on ODI, the participants were divided by quartiles of ODI.

### Fecal metagenomic analysis

SCAPIS participants were instructed to collect fecal samples into dry tubes at home with the provided kit and store the samples at the home freezer until the study site visit. Median interval between fecal sample collection and OSA assessment was 0 days (IQR: 0–1). The samples were then kept at −20°C at the test centers. After 0–7 days, the samples were shipped to the central biobank where they were kept at −80°C. Next, the samples were sent in dry ice to Clinical Microbiomics A/S (Copenhagen, Denmark) for DNA extraction, shotgun metagenomic sequencing, and taxonomic annotation. Analyses were performed in random order of samples’ boxes (16 samples per box) during 2019 and 2020. DNA was extracted from all samples using NucleoSpin® 96 Soil kits (740787; Macherey-Nagel; Germany) from the same batch (Lot: 1903/001). Each extraction round contained a negative and positive control (ZymoBIOMICS^TM^ Microbial Community Standard, D6300, Zymo Research). For cell lysis, a bead beating step was performed for 5 min at 2200 rpm with tubes placed horizontally on the vortex (Vortex-Genie 2).

After DNA fragmentation and library preparation, sequencing was performed using an Illumina Novaseq 6000 system (Illumina, USA). On average, 26.3 million read pairs (SD = ±6.9 millions) were generated per sample for Uppsala samples. Reads were removed if they contained clear adapter contamination, > 10% ambiguous bases, or > 50% bases with Phred quality score < 5. Reads pairs were also removed if any of the reads mapped to the human reference genome GRCh38 using Bowtie 2 (v.02.3.2.)^58^. Reads that passed this quality control were assembled with MEGAHIT (v. 1.1.1)^59^ and mapped using BWA-MEM (v. 0.7.16a)^60^ to a new gene catalog. This new gene catalog was created using samples from the SCAPIS study, which included samples from Uppsala and Malmö sites, samples from the Malmö Offspring Study (MOS)^61^, samples from Pasolli et al.^62^, and 3,486 publicly available genome assemblies from isolated microbial strains.

Co-abundant genes that passed quality assessment were defined as metagenomic species as described in Nielsen et al^49^. Briefly, each metagenomic species has a gene set of 100 signature genes, which are genes selected for optimal and accurate abundance profiling. A metagenomic species was considered detected if read pairs mapped to at least three of the 100 signature genes of that metagenomic species. Then, the total gene counts were used to produce a table of metagenomic species counts, normalized according to read length. Metagenomic species counts were transformed into relative abundances.

There were 1,985 species identified with a mean of 355 species detected per Uppsala sample. Considering the Uppsala participants only, species that were present in ≤1% of the participants were removed, resulting in 1,602 species for subsequent analyses. All the negative controls showed no detectable DNA. DNA extraction from positive controls resulted in a positive signal. For the positive controls, the coefficient of variation estimated by the Shannon diversity index was 3.05%. For 158 pairs of biological replicates, the coefficient of variation was 1.49%. Clinical Microbiomics was unaware that the replicates were submitted for analysis.

The taxonomic annotation of the metagenomic species was performed by mapping the catalog genes to NCBI RefSeq^63^ database (downloaded on May 2, 2021). If >75% of the metagenomic species genes mapped to a single microbial species, a species-level taxonomy was annotated to that metagenomic species. Different thresholds were used for taxonomic annotation at genus, family, order, class, and phylum level (60, 50, 40, 30, and 25% respectively). However, a species or genus-level annotation was not assigned if > 10% of the genes mapped to another single species or genus. For functional annotation, the gene catalog was annotated to the EggNOG (v. 5.0) orthologous groups database (http://eggnogdb.embl.de/) using EggNOG-mapper-software (v. 2.0.1)^64^, which provides annotations to the Kyoto Encyclopedia of Genes and Genomes (KEGG) orthology (KO) database (https://www.genome.jp/kegg/). Based on KO annotations, the metabolic potential of the metagenomic species was determined in terms of GMM^29^. As previously described, the GMMs are 103 metabolic pathways, defined as a series of enzymatic steps represented by KO identifiers^29^. A metagenomic species was considered to contain a GMM if it contained at least two-thirds of the KOs of a module. For modules with three or fewer steps, all steps were required. For modules with alternative paths, only one path had to fulfil the criterion. The relative abundance of a GMM was defined as the sum of the relative abundances of all metagenomic species that encoded that module.

### Anthropometric, sociodemographic, health, and medication information

SCAPIS participants answered an extensive questionnaire on demographic information, education, lifestyle, self-reported health, medication use, as well as a food frequency questionnaire^65^. Smoking was categorized as never, former, or current smoker. Education was categorized based on the highest level achieved. The possible education categories were uncompleted primary or lower secondary education, completed lower secondary education, upper secondary education, or university education. Leisure time physical activity was self-reported as one of four possible categories: mostly sedentary, moderate but regular exercise, moderate activity, regular and moderate activity, or regular exercise or training. According to country of birth, participants were categorized into four groups: born in Scandinavia (Sweden, Denmark, Norway or Finland), in Europe, in Asia, or in other countries. From the food frequency questionnaires, variables were calculated to estimate alcohol intake (g/day)^66^, fiber intake (g/day)^67^, and total energy intake (kcal/day)^66^. Dietary information was assigned as missing for participants whose ln(total energy intake) was greater than the mean of ln(total energy intake) ± 3 standards deviations in the study sample. Anthropometric measurements were performed on site. BMI (kg/m^2^) was calculated as weight divided by the height squared. Systolic and diastolic blood pressure were assessed on site, calculated as the average of two measurements in the arm with the highest mean systolic pressure.

Anti-hypertensive medication and medication for hyperlipidemia were categorized as binary variable based on the questionnaire. Use of proton pump inhibitors (PPI) and use of metformin were defined as a binary variable based on the plasma metabolome. Participants with a measurable omeprazole and/or pantoprazole plasma level were classified as PPI users. Participants with a measurable metformin plasma level were classified as metformin users. Information on previous antibiotic use (Anatomical Therapeutical Chemical code J01) was provided by the SCAPIS cohort study after linkage with the Swedish Prescribed Drug Register (https://www.socialstyrelsen.se/en/statistics-and-data/registers/national-prescribed-drug-register/).

### Plasma metabolome analysis

The fasting plasma samples were collected during site visit and stored at −80°C in the central biobank until they were sent in random order to Metabolon Inc. (Durham, NC, USA) for metabolomics profiling, as previously described^32, 68^. Briefly, after sample preparation, metabolomics was conducted using four protocols: reverse phase (RP)/ultrahigh performance liquid chromatography–tandem mass spectroscopy (UPLC-MS/MS) with negative-ion mode electrospray ionization (ESI), hydrophilic interaction chromatography (HILIC)/UPLC-MS/MS, and two separate RP/UPLC-MS/MS resolutions with positive-ion mode ESI. Metabolites were annotated using Metabolon’s in-house compound library^68^. We used the GUTSY Atlas (https://gutsyatlas.serve.scilifelab.se/) to retrieve information on enrichment for metabolite groups in the gut microbiota species associations with plasma metabolites^32^.

### Statistical analyses

To identify the set of confounders for adjustment, we created a hypothetical causal diagram in the browser-based application DAGitty 3.0 (www.dagitty.net; Fig. S2)^69^. To investigate the association between OSA and the gut microbiota, we identified sex, age, smoking, alcohol intake, and BMI as the minimal sufficient adjustment set. Therefore, these variables were included in the main model together with DNA extraction plate to account for technical variation. Age, alcohol intake, and BMI were modelled as continuous variables. Sex and smoking were modelled as categorical variables. DNA extraction plate was also modelled as a categorical variable with one level per plate (i.e., 52 levels). Given that the hypothetical causal diagram on the effect of OSA on the gut microbiota is rather complex and that confounders such as diet^70^ and socioeconomic status^71^ were not included in the minimal sufficient adjustment set, we chose to construct an extended model accounting for these factors. Thus, the extended model additionally included fiber intake, total energy intake, leisure time physical activity, highest education level achieved, country of birth, and season. Fiber intake and total energy intake were modelled as continuous variables. Leisure time physical activity, highest education level, and country of birth were modelled as categorical variables with four levels each. The variable season consisted of 11 categories based on the month of the study site visit. The categories “June” and “July” were merged because there were only four participants for July.

Analyses were conducted using the R software version 4.1.1 (https://cran.r-project.org/). Partial Spearman’s correlations were performed using the function *pcor.test* in the R package *ppcor*^72^. Gut microbiota beta diversity and alpha diversity were calculated using the package *vegan* version 2.5-7^73^. For the alpha diversity analyses, the Shannon index^22^ was calculated for each participant using the function *diversity*. To investigate how AHI, T90 and ODI were associated with the Shannon index, we used partial Spearman’s correlation. The beta diversity was assessed with the Bray-Curtis dissimilarity, which was calculated using the function *vegdist*. PERMANOVA was conducted using the function *adonis2* (package *vegan*) with 9,999 permutations assessing the marginal effects of the terms. The Bray-Curtis dissimilarity matrix was set as the outcome and the AHI, T90, or ODI severity groups were set as the main exposure separately. Principal coordinate analysis was conducted using the function *pcoa* from the package *ape*^74^.

To investigate how AHI, T90, and ODI associated with the relative abundance of the gut microbiota species, we examined the partial Spearman’s correlation between each of the OSA parameters and each of the 1,602 species. To understand the effect of BMI on our results, we first conducted this analysis with the main model not including the covariate BMI. The species identified in this first step were further investigated in two additional analyses: one with all main model covariates, including BMI, and another model including the extended model covariates. Multiple comparison was accounted for using the Benjamini-Hochberg method with a FDR set at 5%^75^.

The species associated with at least one of the three OSA parameters in the extended model were further examined in three sensitivity analyses. In the first sensitivity analysis, we included to the extended model the variables of medication use, more specifically metformin, PPI, anti-hypertensive medications, and/or medications for hyperlipidemia. In the second sensitivity analysis, we excluded participants (n = 367 among those with valid T90/ODI values) that had used any antibiotic in the previous six months. And in the third sensitivity analysis, we excluded the 29 participants with lung disease, defined as a self-reported diagnosis of COPD, chronic bronchitis, or pulmonary emphysema. After removing these participants, we re-conducted the Spearman’s correlation analyses adjusting for the extended model covariates.

We handled missing data using complete-case analysis; i.e., participants that did not have information for all variables included in a model were removed from that respective analysis. The variable with the highest frequency of missing information was leisure time physical activity followed by smoking, antihypertensive medication use, and use of medication for hyperlipidemia. Of the participants with valid AHI data, 6% were missing information for leisure time physical activity. Information on smoking, anti-hypertensive medication, and medication for hyperlipidemia was missing for 5% of the participants who had valid AHI data. After removing participants with incomplete information on the main model covariates, there were 3,004 participants for AHI analyses and 3,364 participants for the T90 and ODI analyses. Adding or removing BMI did not change the number of participants included, as this variable was not missing for all participants. For the extended model, after removing participants with incomplete information on covariates, there were 2,909 participants for the AHI analyses and 3,249 participants for the T90 and ODI analyses. In the sensitivity analysis adjusting for medication use, there were 3,305 participants for the T90 and ODI analyses.

Because of the lower number of participants with valid AHI values than the number of participants with valid T90 and ODI values, we conducted a secondary analysis where we imputed the AHI values for the 340 participants that for whom we had information on T90 and ODI but not on AHI, considering the extended model covariates. This analysis was conducted using the software Stata 15.1 (Stata Corp., Texas, USA). Multiple imputation was conducted using predicted mean matching with the five nearest neighbors and 10 imputations. Variables used for imputation included all extended model covariates, as well as Shannon index, AHI, T90, ODI, waist-hip-ratio, and the species relative abundance. To avoid including all 1,602 species in the same imputation equation, we performed an imputation step for each species, followed by the Spearman’s correlation analysis between AHI and the species, adjusted for the extended model covariates. The uncertainty of imputations was accounted for using Rubin’s combination rules^76^.

The effect modification by hemoglobin level was explored by categorizing participants into low or high hemoglobin groups based on the sex-specific median hemoglobin level. First, we conducted the Spearman’s correlation of OSA parameters with species relative abundance stratified by hemoglobin group and adjusted for the extended model covariates. Next, each pair of correlation coefficients obtained from the two groups were compared as described in Altman et al.^77^ Briefly, the standard error for each coefficient was estimated using 1,000 bootstrap replications (R package *boot*^78^). Z-scores were then calculated as the ratio of the difference between the two coefficients to the standard error of the difference. The two-sided p-values were then obtained from the standard normal distribution.

For the pathway enrichment analysis for GMM, we used the Spearman’s correlation results from the extended model on the associations of AHI, T90, and ODI with the 1,602 microbiota species. The enrichment analyses were conducted based on ranked p-values using the R package *fgsea*^79^, stratified by the direction of the correlation coefficients. To investigate the metabolite groups associated with each T90/ODI-associated species, we retrieved from GUTSY Atlas^32^ the enrichment analysis results for metabolite groups for each of the species.

In a post hoc analysis, we used Spearman’s correlation to assess the association of our main gut microbiota findings with systolic and diastolic blood pressure, as well as HbA1c. To decide on the set of covariates for adjustment, we created a hypothetical causal diagram of the effect of the gut microbiota on blood pressure and insulin resistance (Fig. S3). Based on the causal diagram, we adjusted the analysis for age, sex, alcohol intake, smoking, fiber intake, total energy intake, leisure time physical activity, country of birth, ODI, T90, and AHI, in addition to DNA extraction plates. Due to the effect of medication on the gut microbiota and the health outcomes of interest, we excluded the participants who self-reported medication use for hypertension from the analyses with blood pressure measurements. In the analyses of the association with HbA1c, we excluded participants who self-reported medication use for diabetes. Using a complete-case analysis, data were available for 2,335 participants for the analyses with blood pressure measurements and for 2,801 participants for the Hb1Ac analyses. Lastly, we assessed the effect of adding BMI to the model.

## Data Availability

The anonymized metagenomic sequences can be found in the European Nucleotide Archive under the accession code "PRJEB51353". The individual-level data underlying this article were provided by the SCAPIS cohort study under agreement, after ethical approval, and are not shared publicly due to confidentiality. Data will be shared upon reasonable request to the corresponding author only after permission by the SCAPIS Data access board (https://www.scapis.org/data-access/) and by the Swedish Ethical Review Authority (https://etikprovningsmyndigheten.se).

## Code availability

The code used in the present analyses is available at GitHub (https://github.com/MolEpicUU/sleepapnea_gut)

## Acknowledgements

We would like to acknowledge the financial support from the European Research Council [ERC-2018-STG801965 (TF); ERC-CoG-2014-649021 (MO-M); ERC-STG-2015-679242 (JGS)], the Swedish Heart-Lung Foundation [Hjärt-Lungfonden, 2019-0505 (TF); 2020-0485 (EL); 2018-0343 (JÄ); 2020-0711 (MO-M); 2020-0173 (GE); 2019-0526 (JGS)], the Swedish Research Council [VR, 2019-01471 (TF), 2018-02784 (MO-M), 2019-01015 (JÄ), 2020-00243 (JÄ), 2019-01236 (GE), 2021-02273 (JGS)], the Swedish Research Council for Sustainable Development [FORMAS, 2020-00989 (SA)], EASD/Novo Nordisk (SA), Göran Gustafsson foundation [2016 (TF)], Axel and Signe Lagerman’s foundation (TF), and governmental funding of clinical research within the Swedish National Health Service (JGS).

We acknowledge the SCAPIS board for enabling the current study, along with the Swedish Heart-Lung Foundation, the main funding body of SCAPIS. Funding for the SCAPIS study was also provided by the Knut and Alice Wallenberg Foundation, the Swedish Research Council and VINNOVA (Sweden’s Innovation agency) the University of Gothenburg and Sahlgrenska University Hospital, Karolinska Institutet and Stockholm County council, Linköping University and University Hospital, Lund University and Skåne University Hospital, Umeå University and University Hospital, Uppsala University and University Hospital. The computations and data handling were enabled by resources in project sens2019512 provided by the Swedish National Infrastructure for Computing (SNIC) at Uppsala Multidisciplinary Center for Advanced Computational Science (UPPMAX), partially funded by the Swedish Research Council through grant agreement no. 2018-05973.

## Competing interests

J.B.H. and H.B.N. are currently working at Clinical Microbiomics A/S. J.Ä. has served on the advisory boards for AstraZeneca and Boehringer Ingelheim and has received lecturing fees from AstraZeneca and Novartis, all unrelated to the present work. C.B. served as a scientific consultant for Repha GmBH, Langenhagen, Germany between 2020 and 2021. JS has stock ownership in companies providing services to Amgen, AstraZeneca, Boehringer, Bayer, Eli Lilly, Itrim, Janssen, Novo Nordisk, Pfizer and Takeda, outside the submitted work. The remaining authors declare no competing interests.

## Contributions

EL, LL, JS, GB, GE, JGS, JÄ, MO-M and TF obtained the funding for the study. GBa, SS-B, JT-H, UH, BK, EL and TF planned and designed the study. JBH and HBN supervised the fecal samples processing and metagenomics bioinformatics analyses at Clinical Microbiomics S/A. GBa carried out the statistical analyses with contribution from SS-B, KD, and UH. GBa wrote first version of the manuscript with support from SS-B, JT-H, BK, EL, and TF. All authors contributed with the critical interpretation of the results and critical revision of the manuscript.

## Supplementary Figures captions

**Figure S1.** Study flowchart for the 4,839 SCAPIS-Uppsala participants with available data on their gut microbiota composition.

**Figure S2.** Directed acyclic graph depicting the causal assumptions on the effect of obstructive sleep apnea (OSA) on the gut microbiota composition. A directed edge (or “arrow”) from one node to another represents a direct effect between these two nodes. Green line: causal path; pink line: biasing path; pink nodes: ancestor of exposure and outcome; and blue nodes: ancestor of outcome.

**Figure S3.** Directed acyclic graph depicting the causal assumptions on the effect of gut microbiota on blood pressure measurements and insulin resistance. A directed edge (or “arrow”) from one node to another represents a direct effect between these two nodes. Green line: causal path; pink line: biasing path; pink nodes: ancestor of exposure and outcome; and blue nodes: ancestor of outcome.

