## Supplementary figures and images for "Obstructive sleep apnea is associated with specific gut microbiota species and functions in the population-based Swedish CardioPulmonary bioImage Study (SCAPIS)"

### Supplementary figure 1

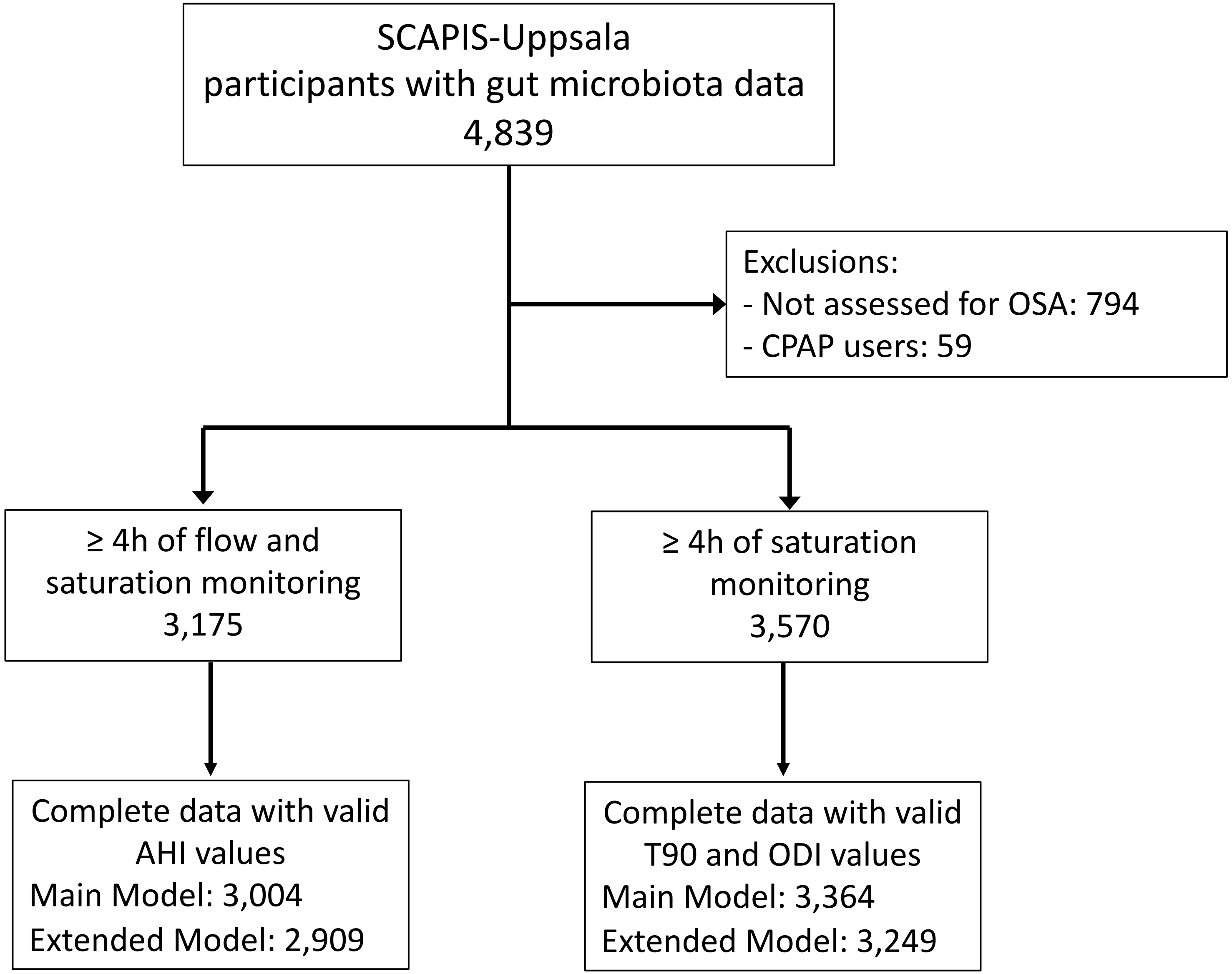

### Supplementary figure 2

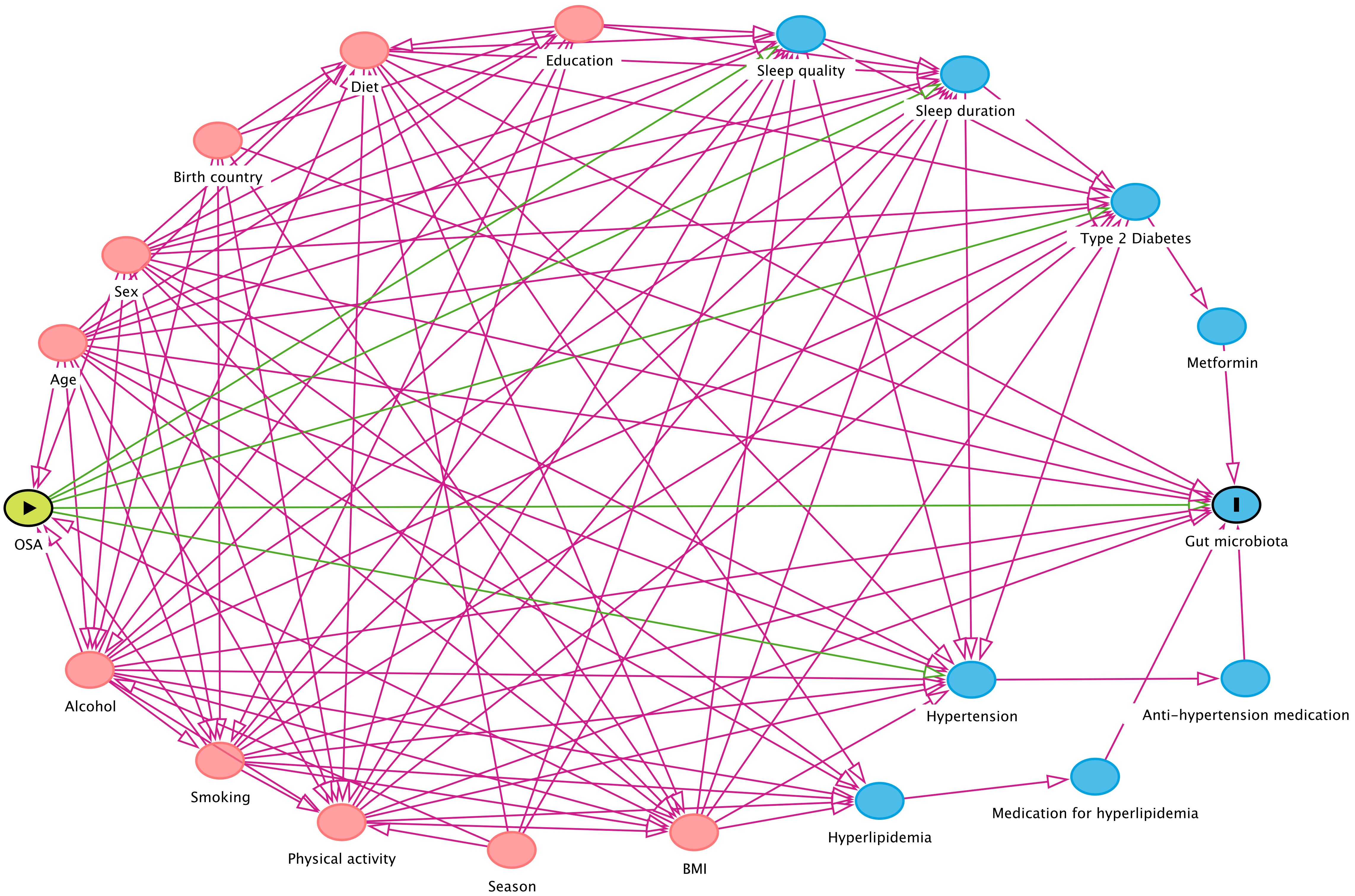

### Supplementary figure 3

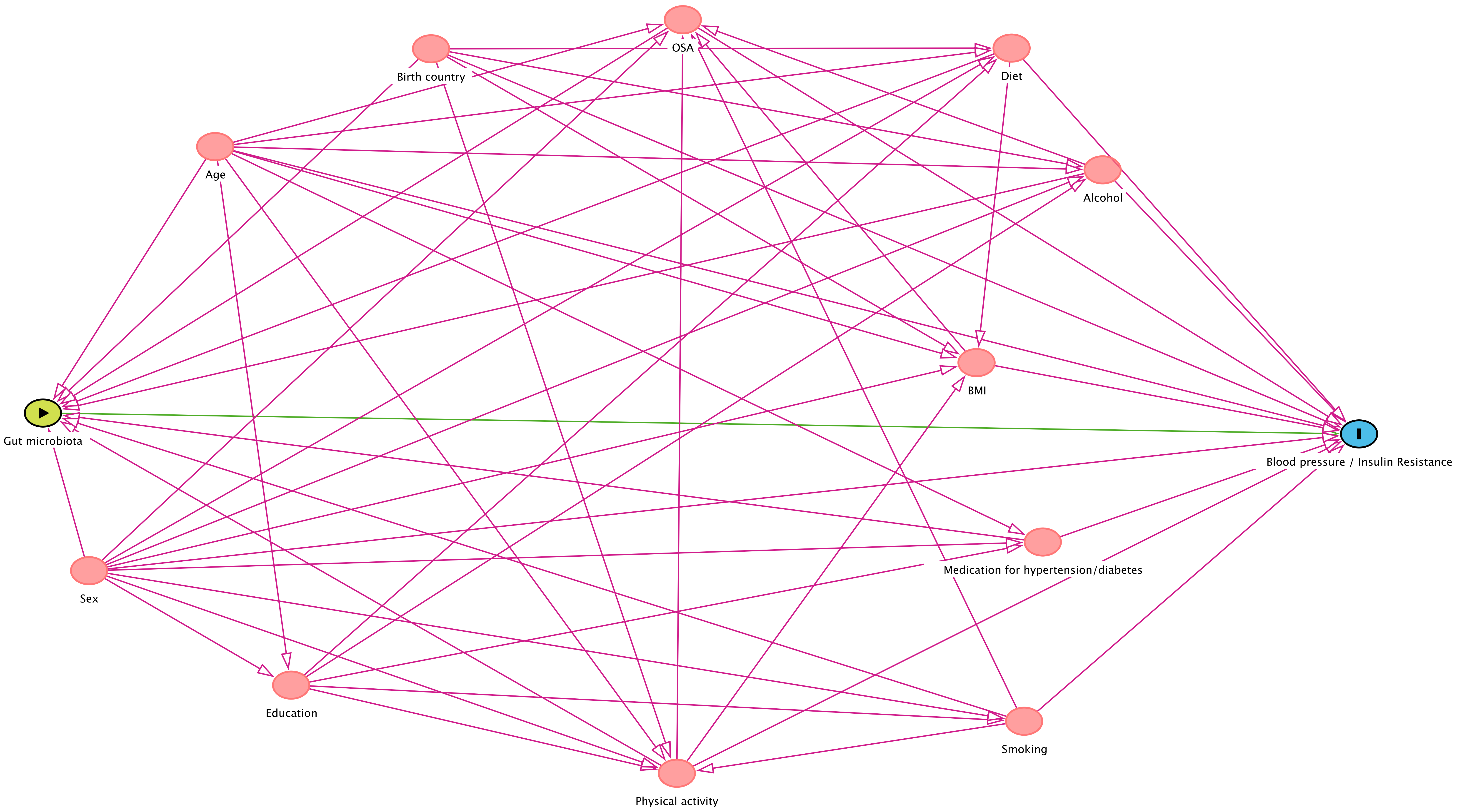
